# Assessment of fractional anisotropy outcomes in combat sport athletes with mild traumatic brain injury

**DOI:** 10.1101/2021.01.06.21249357

**Authors:** Jessica Humara Fonseca, Joe M. Lopez Inguanzo, Janet Perodin Hernández, Evelio González Dalmau

## Abstract

The practice of combat sports increases the risk of suffering white matter injuries. That is why, it is required the early damage detection to determine to what extent the athlete may be active preserving their performance and health status. The integrity of the white matter can be quantitatively characterized in diffusion tensor images, using fractional anisotropy. This study aims at characterizing the fractional anisotropy of white matter injuries in combat athletes that are exposed to repetitive trauma and also, to detect changes in fractional anisotropy between cerebral hemispheres with and without lesions. It is proposed a global and structural analysis of the hemispheres, as well as the selection of ROI in the lesions. 14 athletes, from Boxing, Karate and Taekwondo sports, participated. The sample was divided into two groups of seven subjects each: Injured (23.428±4.157 years old) and Healthy (24.285±5.023 years old) paired by sport denomination. Diffusion tensor images were used to obtain FA values in the analysis of the hemispheres and lesions. Global and structural analysis of the hemispheres did not detect the presence of white matter lesions; however, the use of ROI selection permitted maximum approximation of the injuries location. It also improved the breakdown of FA values as it allows a local analysis of the lesion. As an additional result, there were found ROIs values, FA_*med*_ = 0.454±0.062, which exceed the average fractional anisotropy of the white matter. The cohesion of acute and chronic phase lesions were found in the same subject. The apparently contradictory results in FA values are related to the stage of the lesions.

## 1 Introduction

The practice of combat sports increases the risk of suffering white matter injuries (WMI), which modifies the pattern of diffusion of the affected area (Churchill et al., 2017b). Eighty percent of white matter injuries are classified as mild traumatic brain injuries (mTBI) (Langlois et al., 2006). The integrity of the white matter can be quantitatively characterized by diffusion tensor images (DTI) (King et al., 2019). This technique provides information on the diffusion of water molecules from different metrics usage such as fractional anisotropy, mean diffusivity, Axial diffusivity, and radial diffusivity. The fractional anisotropy (FA), which values vary between zero and one, is one of the most used parameters to characterize white matter lesions (Chang et al., 2017; Wilde et al., 2015). An FA coefficient close to one, occurs, when water molecules diffuse in a restricted medium like in the presence of membranes, filaments and myelin sheaths (anisotropic diffusion). In contrast, an FA coefficient close to zero, is observed, in the cerebrospinal fluid, where the water molecules diffuse in all directions (isotropic diffusion) with the same probability and speed (Koerte et al., 2015).

There is a contradiction in the consulted literature regarding the reported values of FA in subjects with white matter lesions. Some authors reported higher values of FA in sport injured athlete, rather than in sport healthy control athlete; while others revealed lower FA in sport injured athlete in comparison to non-athlete controls (Asken et al., 2018). The reason for conflicting diffusion direction (increased vs. decreased anisotropy) acutely/subacutely after sport-related concussion across studies is not entirely clear (Brett et al., 2019). We hypothesized that the apparent contradiction is caused by the coherence between the acute phase injuries and chronic phase ones. The objective of this study is to characterize clinically asymptomatic injuries in combat athletes (Boxing, Karate and Taekwondo) exposed to repetitive trauma. We also intend to detect changes in FA values between cerebral hemispheres with and without lesions. It is proposed a global and structural analysis of the hemispheres, as well as the selection of ROI in the lesions.

## 2 Material AND Methods

### 2.1 Participants

In this study participated male athletes from combat sports: Boxing, Karate and Taekwondo with ages ranging from 19 to 35 years old. The sample was taken from the data base of the Cuban National Sport Institute (INDER) and selected under the criteria of being athletes of high performance and the positions of their cranial injuries during competition and training. Those with claustrophobia and symptoms of neurological disorders were excluded. As a result of the inclusion and exclusion criteria, 14 subjects were selected; from them, 13 active athletes and one coach. The sample was divided into two groups of seven subjects each: Injured (23.428 ± 4.157 years old) and Healthy (24.285 ±5.023 years old) paired by sport denomination (Supplementary Material Table [S1]).

### 2.2 Image acquisition

Allegra 3T Magnetic Resonance Imaging Equipment (Siemens) was used to locate the white matter lesions. Structural imaging included T1-weighted images (*TR* = 2000*ms, TE* = 2.6*ms* and slice thickness=1*mm*), T2-weighted images (*TR* = 3500*ms, TE* = 354*ms* and slice thickness=1.20*mm*), *T* 2^*^ susceptibility-weighted images to rule out microbleeds (*TR* = 656*ms, TE* = 20*ms* and slice thickness 2*mm*).*TR* = 656*ms, TE* = 20*ms* and slice thickness 2*mm* and FLAIR imaging (*TR* = 5000*ms, TE* = 353*ms* and slice thickness 1.20*mm*). A diffusion-weighted imaging protocol was performed (*TR* = 10900*ms, TE* = 94*ms*, resolution 2*mm* and slice thickness=2.3*mm*) consisting of 61 diffusion-weighting directions, *b* = 1000*s*/*mm*^2^ and *b* = 0 scans.

### 2.3 Image Processing and Data Analysis

Diffusion-weighted images (DWIs) were processed to obtain Diffusion Tensor Images (DTI) using DSI-Studio software (http://dsi-studio.labsolver.org). During the process, distortion corrections and co-registration were performed with FLAIR images. Four quantitative diffusion descriptors were obtained from the resulting images: fractional anisotropy, mean diffusivity, axial diffusivity and radial diffusivity. The present study only focused on fractional anisotropy. For data processing, applying DTI and subsequent statistical analyzes, the sample was divided into two groups: healthy and injured of seven subjects each. Additionally, the accuracy index was calculated in the global and structural analyzes. To determine how the mean fractional anisotropy FA_*med*_ varies in the hemispheres with the presence of lesions, we obtained 14 values of FA for each of the three methods of analysis: global, structural and ROIs. In the first case, atlases of the hemispheres and lateral ventricles were used (Hutchinson et al., 2018; Kikinis et al., 2017). The FA cerebrospinal fluid (CSF) present in the lateral ventricles and, the FA in the white matter (WM) of the hemispheres of subjects without lesions, were taken as a reference of maximum and minimum diffusion. In the structural analyzes, the procedure is similar to the previous one regarding the use of atlases of the structures studied (McAllister et al., 2014). Those with the highest density of tracts and/or length were used as selection criteria. As a result of this criterion, eight structures were selected: Genu of Corpus Callosum (GCC) (Polak et al., 2015), Splenium of Corpus Callosum (SGG), Anterior Corona Radiata (ACR), Posterior Corona Radiata (PCR) and Superior Corona Radiata (SCR) (Dean et al., 2015). Anterior Limb of Internal Capsule (ALIC) (Dean et al., 2015), Inferior Fronto-Occipital Fasciculos (IFOF) and Superior Longitudinal Fasciculos (SLF) (Zivadinov et al., 2018). The study of hemispheres were done by manually marked ROIs of the lesions (Zhang et al., 2006). As a classifier of the behavior of the ROIs, there were used the values of FA_*med*_ of the cerebrospinal fluid and the white matter of subjects with lesions analysis. OriginPro 8 software was used to calculate the mean and standard deviations of the FA, besides the confidence intervals for the mean in the FA (95% confidence). Finally, t-student tests were performed to compare the results obtained for each hemispheres in the techniques used.

## 3 Results

*FLAIR* and *T* 2 images identified 18 white matter lesions and they were recorded by their hyperintense contrast Fig. 1. In general, the right hemisphere is more occupied with lesions than the left one. Diffusion tensor images were obtained to quantify FA in the hemispheres and in the lesions, utilizing global analyzes by structure and by selection of ROIs.

**Figure 1:**
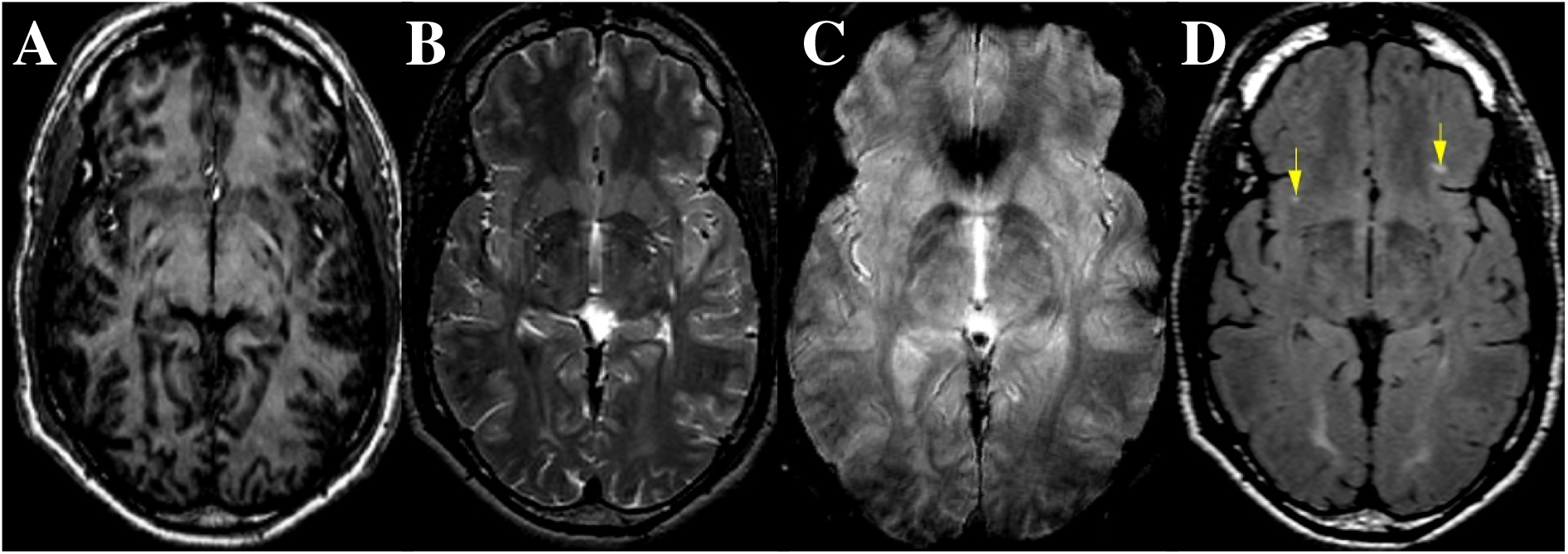
Images of the subject CN0946-18. T1-weighted images **(A)**, T2-weighted images **(B)**, *T* 2^*^ susceptibility-weighted images **(C)** and FLAIR imaging **(D)**. Yellow arrows point the lesions on FLAIR image.

The global analyzes, Fig. 2**A**, revealed maximum values of FA_*med*_(*Healthy*) = 0.403± 0.007 while the lowest values were reported in the cerebrospinal fluid FA_*med*_(*In jured*) = 0.163 ±0.037, Table 1. No significant differences were detected between the hemispheres of subjects with and without injuries. There were either statistically significant differences between hemispheres of the both group Table 1. This method presents an accuracy index=0.479.

**Table 1:** Global analyzes in the hemispheres of healthy subjects and subjects with white matter lesions.

| Global Analysis | Injured ($FA_{med}$) | | | Healthy ($FA_{med}$) | | | p value | |
| --- | --- | --- | --- | --- | --- | --- | --- | --- |
| | Left | Right | $p^*$ | Left | Right | $p^*$ | Left | Right |
| White Mater | $0.396 \pm 0.015$ | $0.396 \pm 0.017$ | 0.937 | $0.400 \pm 0.006$ | $0.403 \pm 0.007$ | 0.528 | 0.578 | 0.368 |
| Cerebrospinal Fluid | $0.176 \pm 0.031$ | $0.163 \pm 0.037$ | 0.486 | $0.184 \pm 0.054$ | $0.180 \pm 0.050$ | 0.893 | 0.751 | 0.487 |

$p^*$ :t-student score between right and left hemispheres.
**p value**:t-student score between injured and healthy subjects.

**Figure 2:**
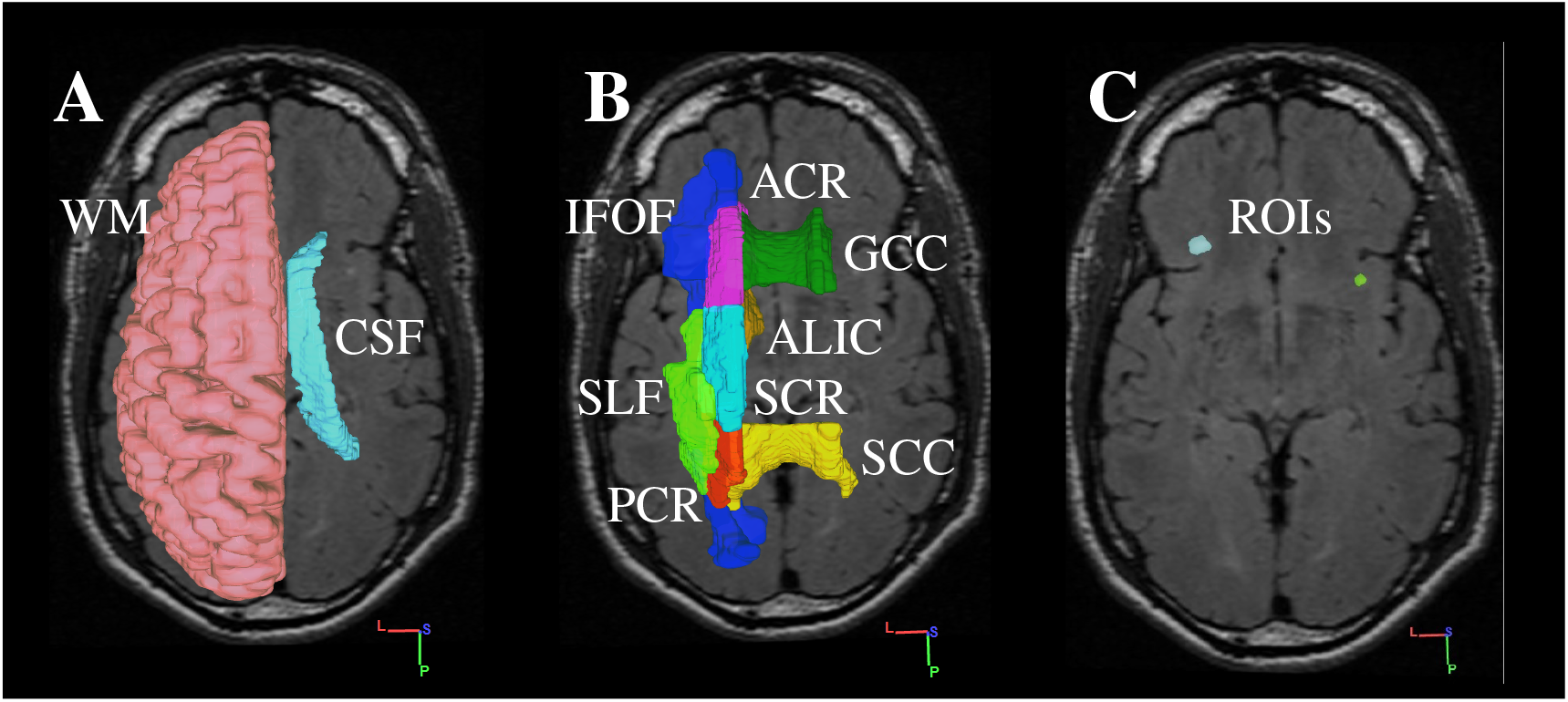
Analysis of the hemispheres with lesions. In the global analyzes (A) the cerebrospinal fluid (CSF) present in the lateral ventricles and white matter (WM). The Genu of Corpus Callosum (GCC), Splenium of Corpus Callosum (SGG), Anterior Corona Radiata (ACR), Posterior (PCR), Superior (SCR) Anterior Limb of Internal Capsule (ALIC), Inferior Fronto-Occipital Fasciculos (IFOF) and Superior Longitudinal Fasciculos (SLF) used for the analysis by structures (B).(C) Analysis of Regions of Interest (ROIs).

The values of FA_*med*_ in the analyzed structures, Fig. 2**B**, had higher mean values than those obtained in the global analyzes Table 2, consistently with the largest organization of white matter in structures. The highest values of FA_*med*_(*Healthy*) = 0.678± 0.019 are found in the SCC and the lowest FA_*med*_(*In jured*) = 0.392 ± 0.021 is located in the left IFOF. Structural analyzes do not detect differences between hemispheres with and without injuries; except in the case of the ACR Table 2. This method presents an accuracy index_*m*_*ed* = 0.396.

**Table 2:**
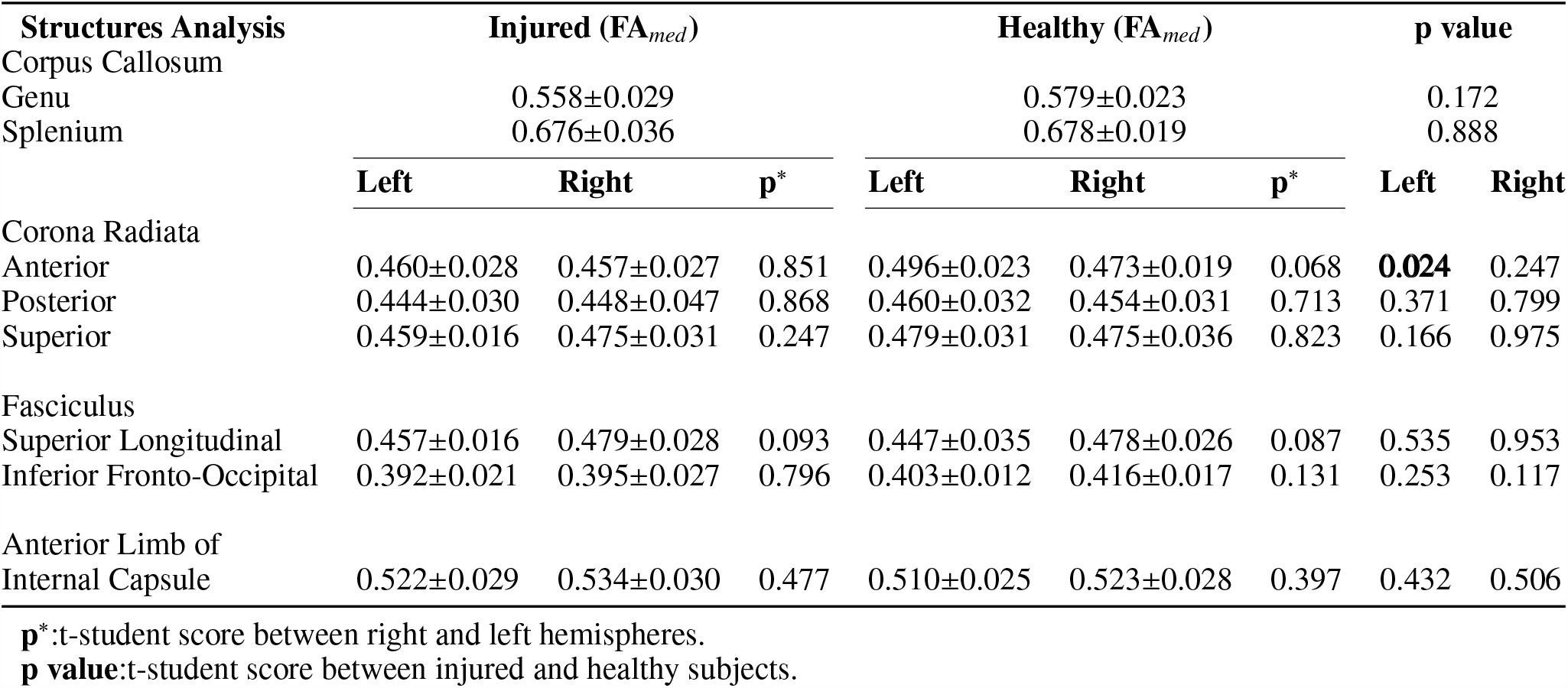
Structural analyses in the hemispheres of healthy subjects and subjects with white matter lesions.

| Structures Analysis | Injured ($FA_{med}$) | | | Healthy ($FA_{med}$) | | | p value | |
| --- | --- | --- | --- | --- | --- | --- | --- | --- |
| Corpus Callosum |  |  |  |  |  |  |  |  |
| Genu | | $0.558 \pm 0.029$ | | | $0.579 \pm 0.023$ | | | 0.172 |
| Splenium | | $0.676 \pm 0.036$ | | | $0.678 \pm 0.019$ | | | 0.888 |
| | Left | Right | $p^*$ | Left | Right | $p^*$ | Left | Right |
| Corona Radiata |  |  |  |  |  |  |  |  |
| Anterior | $0.460 \pm 0.028$ | $0.457 \pm 0.027$ | 0.851 | $0.496 \pm 0.023$ | $0.473 \pm 0.019$ | 0.068 | **0.024** | 0.247 |
| Posterior | $0.444 \pm 0.030$ | $0.448 \pm 0.047$ | 0.868 | $0.460 \pm 0.032$ | $0.454 \pm 0.031$ | 0.713 | 0.371 | 0.799 |
| Superior | $0.459 \pm 0.016$ | $0.475 \pm 0.031$ | 0.247 | $0.479 \pm 0.031$ | $0.475 \pm 0.036$ | 0.823 | 0.166 | 0.975 |
| Fasciculus |  |  |  |  |  |  |  |  |
| Superior Longitudinal | $0.457 \pm 0.016$ | $0.479 \pm 0.028$ | 0.093 | $0.447 \pm 0.035$ | $0.478 \pm 0.026$ | 0.087 | 0.535 | 0.953 |
| Inferior Fronto-Occipital | $0.392 \pm 0.021$ | $0.395 \pm 0.027$ | 0.796 | $0.403 \pm 0.012$ | $0.416 \pm 0.017$ | 0.131 | 0.253 | 0.117 |
| Anterior Limb of Internal Capsule | $0.522 \pm 0.029$ | $0.534 \pm 0.030$ | 0.477 | $0.510 \pm 0.025$ | $0.523 \pm 0.028$ | 0.397 | 0.432 | 0.506 |

$p^*$ :t-student score between right and left hemispheres.
**p value**:t-student score between injured and healthy subjects.

Manually selected ROIs measurements were done on the diffusion maps Fig. 2**C**. 18 ROIs of the lesions, which mean volume is 0.011cc were marked. From those, 14 with statistical values of FA_*med*_ = 0.271±0.057 similar to the cerebrospinal fluid. In contrast, four ROIs with FA_*med*_ = 0.454 ± 0.062 were detected. Both types of ROI, with significant FA value differences of **p**=8.829× 10^−13^, coexist in three subjects.

These ROIs marked by a *Star Symbol* Table 3 were classified as atypical since they presented FA values that exceed the FA_*med*_ (Injured) of the white matter. The mean volumes of the structural ROIs, hemispheres, ventricles and ROIs of lesions were calculated using the three techniques (Supplementary Material Table [S2]). The same procedures were performed to quantify the FA confidence intervals (Supplementary Material Table [S3]). Finally, it was compared the white matter and cerebrospinal fluid FA values of subjects with lesions, including the two types of ROIs found, Fig. 3.

**Table 3:**
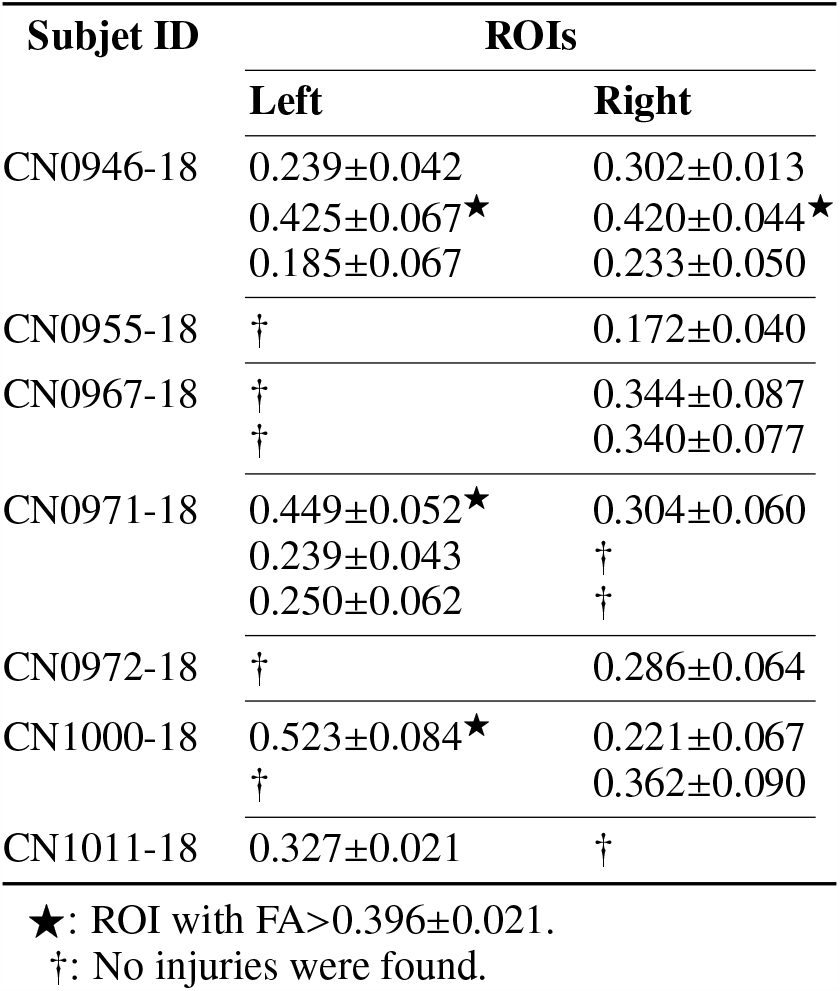
Fractional anisotropy in ROIs of lesions.

**Figure 3:**
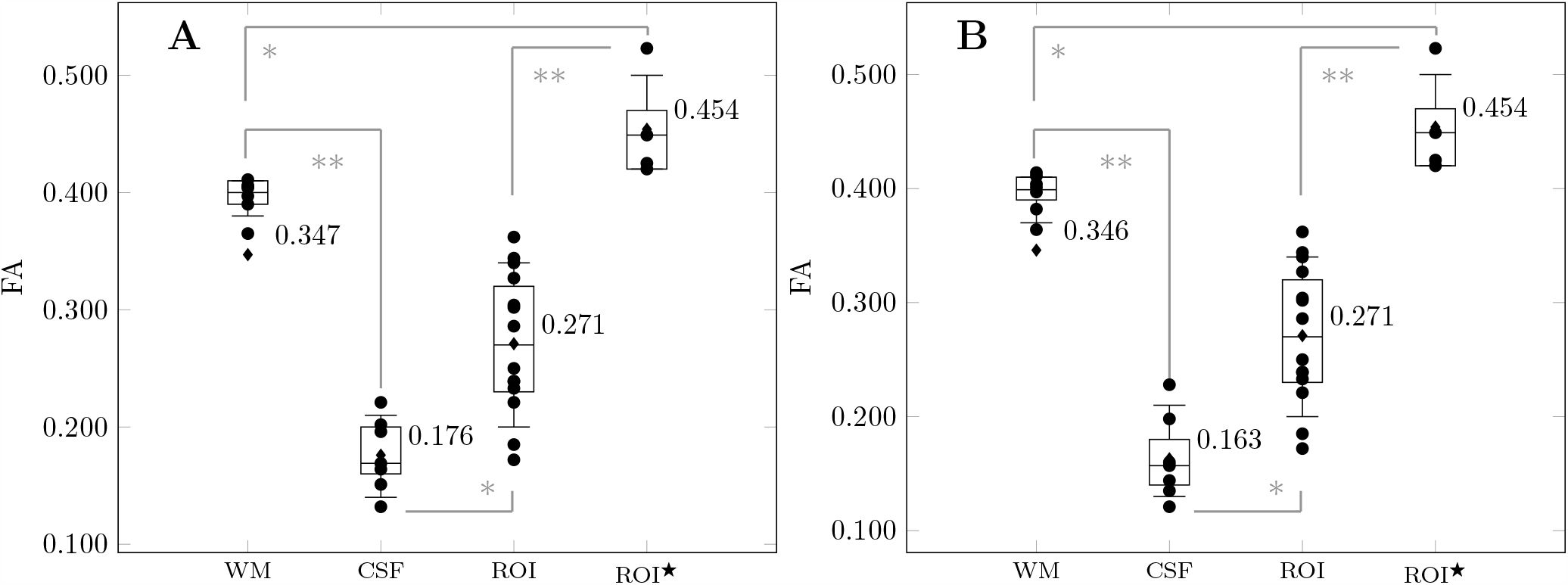
Comparison of FA values in injured subjects on White Matter, Cerebroespinal fluid and ROIs in left (A) and right (B) hemispheres. *Star Symbol* represents the ROIs with FA>WM). Significant differences (*p*^*^ < 0.005; *p*^**^ < 0.001).

## 4 Discussion

The present study demonstrates that the global and structural analyzes do not identify differences in the FA_*med*_ values of the injured hemispheres, despite the existence of marked variances between both injured hemispheres. The uneven distribution of injuries is either detected when comparing the means of the FA between the hemispheres of injured subjects with respect to healthy subjects.

The global analysis of the hemispheres did not result in a sensible test since the average volume of the lesions represents 0.005% (Supplementary Material Table [S2]) of the right average volume and, 0.006% of the left average volume of the white matter in injured hemispheres. This fact masks differences between subjects with and without lesions as the volume of the lesions is negligible in respect to the volume of the hemispheres. Therefore, the values of FA in lesions do not significantly contribute to the global FA mean in the hemispheres, unlike the results found by Kikinis Z (Kikinis et al., 2017), who detected considerable lesion volume with respect to the volume of the hemisphere in animals with recent lesions. However, Zhang L (Zhang et al., 2006) found that after a month of sustaining a concussion, injured athletes did not differ from non-concussed athlete controls FA on the whole-brain.

Although the structural analyzes allow a greater approach to the location of the lesions, it does not constitute a sensitive test since lesions are asymptomatic; so, the location of the possible damaged structures are exactly unknown. In the studied samples, the impacts were in different areas, thus the position of the lesions is ‘unique’ in each subject. In the ACR selected structures, there were noteworthy differences between healthy and injured subjects. SLF showed higher FA_*med*_ (injured) in the right hemisphere while Meier T (Meier et al., 2016) reported an increase of the FA_*med*_ in the left hemisphere. The results of the GCC, FA_*med*_(*in jured* < *Healthy*) contradicts what was found by Ware (Ware et al., 2020). Lastly, in the ACR FA_*med*_(*in jured* < *Healthy*) in both hemispheres, resulted similar to the ones stated by Yin B (Yin et al., 2019); he only reported a decrease of the FA values in the left hemispheres of injured subjects, though. In all structures, the percentage that represents the volume of the lesions is greater than in the case of the global analyzes (Supplementary Material Table [S2]), so that the contribution of the lesions to the FA increases.

In global and structural techniques, it is observed that the presence of lesions increases the coefficients of variation of the FA (Supplementary Material Table [S3]). This implies that the trends of the mean do not change but their deviation standard. Therefore, global and structural analyzes detect the presence of lesions based on variations in the standard deviation. These analyzes are not reliable as the unique criterion for determining the presence or absence of asymptomatic lesions, due to the unawareness of occurrence time and impact zone. In those tests, accuracy index below 0.5 were obtained, showing its low sensitivity to the presence of lesions (Supplementary Material Table [S4]).

Unlike the previous techniques, with the selection of ROIs, a maximum approximation for the location of the lesions is obtained. As a further result, ROIs were found with atypical FA values that exceed the FA_*med*_ of the white matter. Several authors have observed this pattern in the first days after a mild traumatic brain injury occurred (Churchill et al., 2017a; Wallace et al., 2018). No relationship was found between atypical FA values and the volume of the selected ROIs. Other authors reported that FA follows a decreasing trend in subjects with lesions of four years or more (Niogi et al., 2008). Figure 3 shows lesions in the two phases of their temporal evolution. The acute phase, lesions manifest FA values, which exceed the mean white matter FA, herein indicated with a *Star Symbol*. The presence of chronic phase lesions is evident, which FA values is similar to the media values of the cerebrospinal fluid. This suggests that the injuries were made at different times and therefore, their evolution is at different stages. According to that, the apparent contradiction with respect to the FA values in the white matter referred by Asken B. and Brett (Asken et al., 2018; Brett et al., 2019) are solved. These findings should be added to the list of factors involved in the conflict diffusion direction (increased vs. decreased anisotropy) acutely/subacutely mentioned by Dodd AB (Dodd et al., 2014).

## 5 Conclusion

In the consulted literature, the apparently contradictory results in the FA values are related to the stage of the lesions. The reported analysis in the referred studies are either global, structures or selection of ROIs, rather than using the three methods simultaneously. The present study found that there were not previous report of both lesions (acute and/or chronic) in the same subject. In spite of our knowledge that there is evidence of injures in the studied subjects, the global and structural analyzes mask those results; therefore, we do not recommend these types of analysis for determining the existence of lesions. Global and structural analyzes should be used as a complement to the MR images when the presence of lesions, their location and/or the time elapsed is not known a priori. It has been demonstrated that, the best criterion for the analysis of the hemispheres with lesions, is the selection of ROIs lesions, as it permits a better location of the lesion and improves the estimation of FA values, allowing a local analysis of the lesion and the concurrence of acute and chronic phase lesions cohesion in the same subject.

### Limitation of the study

Although, the current investigation does not include a wide sample of the exploratory groups and the heterogeneity of age range, it establishes the scientific basis for a further and detailed analysis on this topic. Psychological and social evaluation of subjects, object of study, would be considered.

## Data Availability

The [GENERATED / ANALYZED] data sets for this study can be requested at

## Conflict of Interest Statement

The authors declared that they had no conflicts of interest with respect to their authorship or the publication of this article. No competing financial interests exist.

## Author Contributions

All authors contributed intellectually to manuscript revision, read, and approved the submitted version.

## Acknowledgments

Authors would like to thank the considerations and recommendations to the present study, of Professor and Ph.D. Maria Antonieta Bobes, from the Department of Cognitive Neuroscience of the Cuban Neuroscience Center. The authors would like to thank the Cuban National Sport Institute (INDER).

## Data Availability Statement

The [GENERATED / ANALYZED] data sets for this study can be requested at.

---

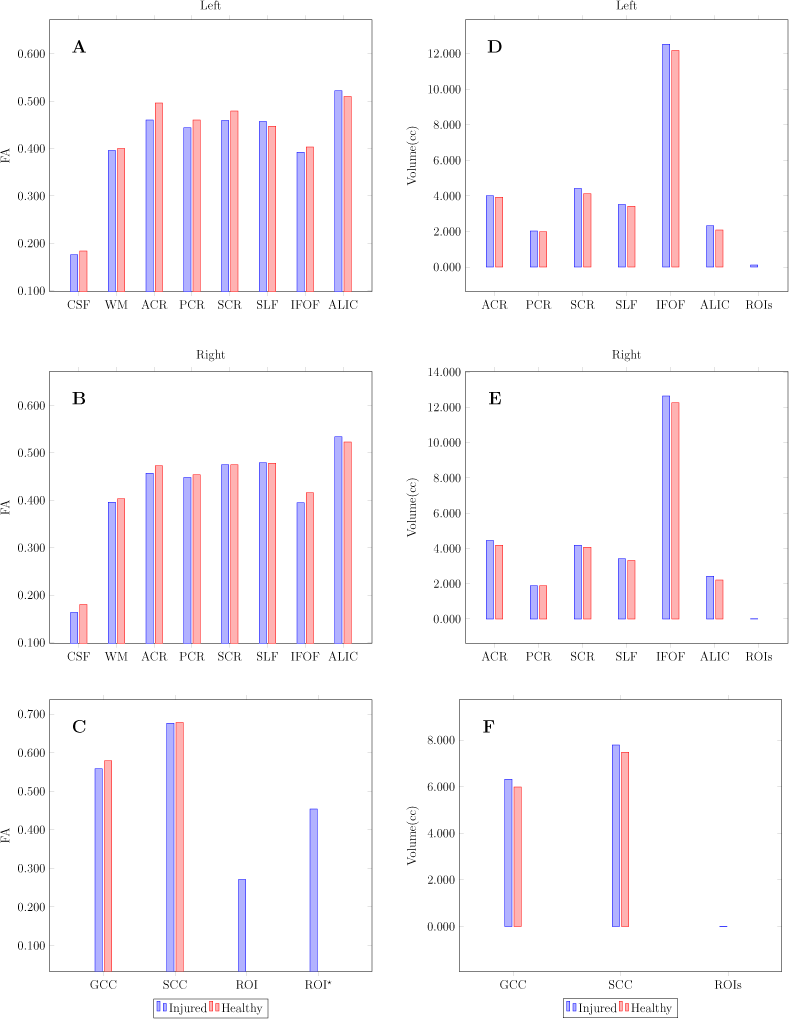

## Notes

### Competing Interest Statement

The authors have declared no competing interest.

### Author Declarations

Approved by the ethics committee of the Center for Neurosciences of Cuba

