## Additional tables and figures for "Assessment of fractional anisotropy outcomes in combat sport athletes with mild traumatic brain injury"

Preprint, compiled December 28, 2020

### 1 Supplementary Tables and Figures

Table 1: Subject demographic characteristics.

| Subject ID | Injured | | | Subject ID | Healthy | | |
|---|---|---|---|---|---|---|---|
| CN0946-18 | BOXING | ACTIVE | 25 | CN0977-18 | BOXING | ACTIVE | 24 |
| CN0955-18 | KARATE | ACTIVE | 19 | CN0953-18 | KARATE | ACTIVE | 20 |
| CN1000-18 | KARATE | ACTIVE | 30 | CN0954-18 | KARATE | ACTIVE | 23 |
| CN0967-18 | JUDO | ACTIVE | 21 | CN0944-18 | JUDO | ACTIVE | 21 |
| CN0971-18 | JUDO | COACH | 27 | CN0969-18 | JUDO | ACTIVE | 25 |
| CN0972-18 | JUDO | ACTIVE | 19 | CN0968-18 | JUDO | ACTIVE | 22 |
| CN1011-18 | JUDO | ACTIVE | 23 | CN0976-18 | JUDO | ACTIVE | 35 |

(BOXING): 2 (KARATE): 4 (JUDO): 8
(ID): Personal indicator

Table 2: Mean, standard deviation and coefficients of variation in volumes.

| Volume(cc) | Injured | | | | Healthy | | | |
|---|---|---|---|---|---|---|---|---|
| Corpus Callosum | | | | | | | | |
| Genu | 6.315±0.783 | 0.124 | | | 5.985±0.651 | 0.108 | | |
| Splenium | 7.788±0.711 | 0.091 | | | 7.477 ±1.166 | 0.156 | | |
| ROIs$_{med}$ | 0.011±0.012 | 1.088 | | | | | | |
| | **Left** | **C$_v$** | **Right** | **C$_v$** | **Left** | **C$_v$** | **Right** | **C$_v$** |
| White Mater | 186.068±18.713 | 0.100 | 181.665±18.779 | 0.103 | 169.820±20.229 | 0.119 | 165.995±20.646 | 0.124 |
| Cerebrospinal Fluid | 4.916±1.469 | 0.298 | 4.237±1.280 | 0.302 | 5.021±0.624 | 0.124 | 4.234±0.504 | 0.119 |
| Corona Radiata | | | | | | | | |
| Anterior | 4.007±0.466 | 0.116 | 4.448±0.485 | 0.109 | 3.924±0.397 | 0.101 | 4.179±0.367 | 0.087 |
| Posterior | 2.030±0.189 | 0.09 | 1.881±0.241 | 0.128 | 1.984±0.320 | 0.161 | 1.881±0.345 | 0.183 |
| Superior | 4.414±0.622 | 0.140 | 4.185±0.487 | 0.116 | 4.119±0.551 | 0.133 | 4.062±0.648 | 0.159 |
| Fasciculus | | | | | | | | |
| Superior Longitudinal | 3.519±0.394 | 0.112 | 3.417±0.365 | 0.107 | 3.422±0.650 | 0.189 | 3.311±0.555 | 0.167 |
| Inferior Fronto-Occipital | 12.527±1.091 | 0.087 | 12.643±1.231 | 0.097 | 12.180±1.988 | 0.163 | 12.265±1.569 | 0.127 |
| Anterior Limb of Internal Capsule | 2.324±0.376 | 0.161 | 2.428±0.303 | 0.125 | 2.084±0.290 | 0.139 | 2.206±0.213 | 0.096 |

**C$_v$**: variation coefficient

Publication doi

Preprint doi

Table 3: Confidence intervals for the mean of the fractional anisotropy measurements (95% confidence).

| | **Injured** | | | | **Healthy** | | | |
|---|---|---|---|---|---|---|---|---|
| Corpus Callosum | | | | | | | | |
| Genu | [0.507;0.561] | 0.090 | | | [0.566;0.594] | 0.044 | | |
| Splenium | [0.641;0.681] | 0.054 | | | [0.656;0.692] | 0.048 | | |
| ROI | [0.246;0.309] | 0.206 | | | | | | |
| ROI$^\star$ | [0.441;0.510] | 0.131 | | | | | | |
| | **Left** | **C$_v$** | **Right** | **C$_v$** | **Left** | **C$_v$** | **Right** | **C$_v$** |
| White Mater | [0.380;0.404] | 0.054 | [0.378;0.402] | 0.054 | [0.392;0.408] | 0.039 | [0.393;0.411] | 0.034 |
| Cerebrospinal Fluid | [0.137;0.178] | 0.236 | [0.131;0.171] | 0.239 | [0.159;0.263] | 0.440 | [0.152;0.254] | 0.453 |
| Corona Radiata | | | | | | | | |
| Anterior | [0.429;0.467] | 0.079 | [0.412;0.460] | 0.101 | [0.479;0.505] | 0.074 | [0.465;0.487] | 0.044 |
| Posterior | [0.411;0.449] | 0.080 | [0.422;0.468] | 0.093 | [0.439;0.467] | 0.058 | [0.439;0.475] | 0.072 |
| Superior | [0.435;0.463] | 0.058 | [0.435;0.479] | 0.086 | [0.462;0.492] | 0.057 | [0.456;0.492] | 0.070 |
| Fasciculus | | | | | | | | |
| Superior Longitudinal | [0.438;0.474] | 0.071 | [0.452;0.493] | 0.078 | [0.435;0.475] | 0.080 | [0.462;0.512] | 0.093 |
| Inferior Fronto-Occipital | [0.377;0.403] | 0.060 | [0.381;0.409] | 0.064 | [0.394;0.410] | 0.037 | [0.400;0.422] | 0.049 |
| Anterior Limb of Internal Capsule | [0.503;0.539] | 0.061 | [0.521;0.559] | 0.066 | [0.500;0.532] | 0.057 | [0.514;0.542] | 0.048 |

**C$_v$**: variation coefficient

Table 4: Calculation of the accuracy indices in global and structural analyzes.

| Analysis | | $acc = \frac{TP+TN}{TP+FP+TN+FN}$ |
|---|---|---|
| Global | Hemispheres | 0.479 |
| Structures | | |
| | Genu | 0.326 |
| | Splenium | 0.530 |
| | SCR | 0.295 |
| | ACR | 0.183 |
| | PCR | 0.398 |
| | SLF | 0.520 |
| | IFOF | 0.326 |
| | ALIC | 0.591 |

Figure 1: Comparison of $FA_{med}$ of the right **(A)** and the left **(B)** hemispheres through global and structural analyzes, as well as the assessment of the Corpus Callosum (GCC and SCC) the ROI and ROI$^{\star}$ **(C)**. Comparison of $Volumenes_{med}$ between the right **(D)** and the left **(E)** hemispheres in the analysis by structures, as well as the one of the Corpus callosum and the ROIs **(F)**.

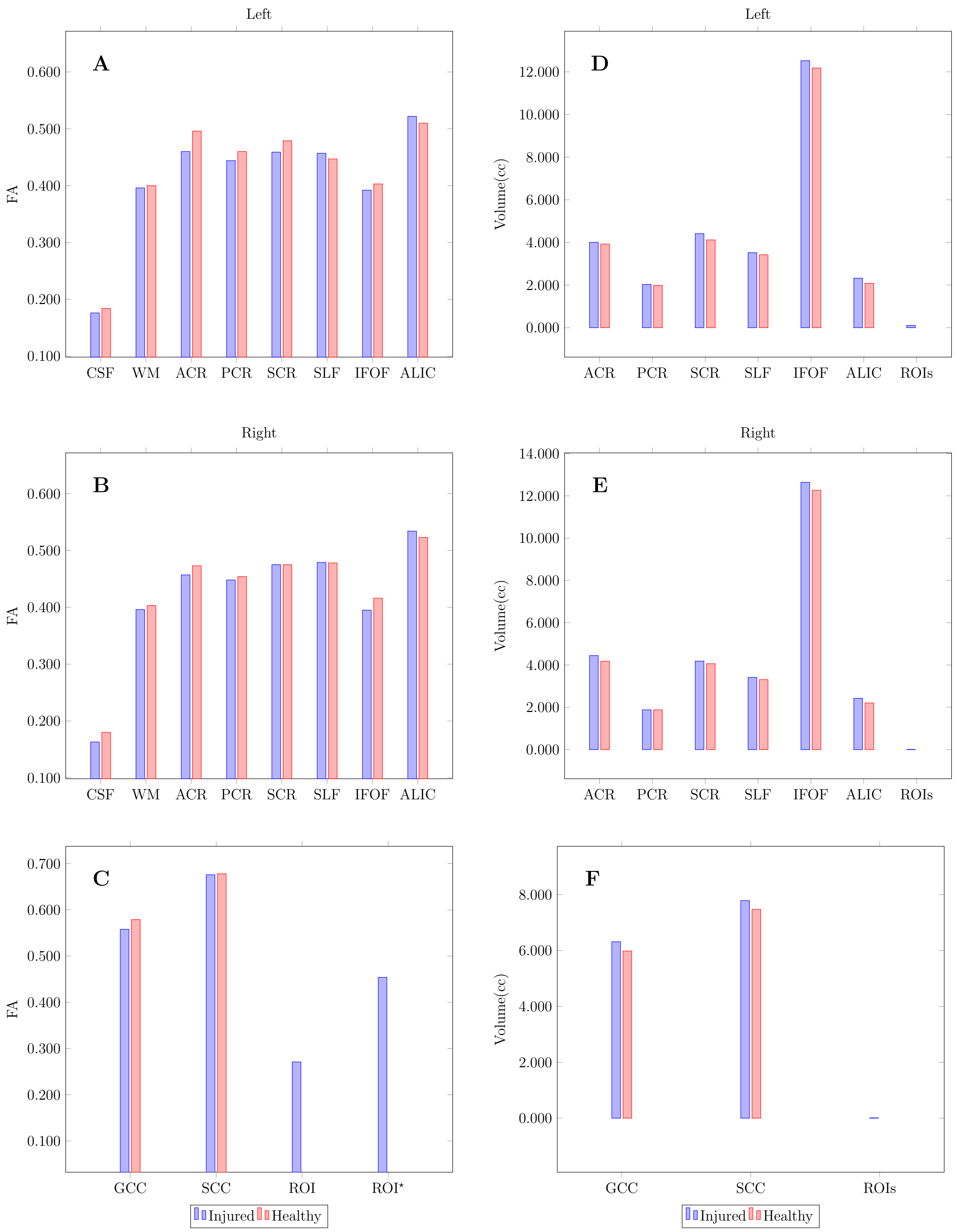
